# Estimating the Effect and Cost-Effectiveness of Facemasks in Reducing the Spread of the Severe Acute Respiratory Syndrome-Coronavirus 2 (SARS-CoV-2) in Uganda

**DOI:** 10.1101/2020.06.11.20128272

**Authors:** Betty K Nannyonga, Rhoda K Wanyenze, Pontiano Kaleebu, John M Ssenkusu, Tom Lutalo, Fredrick Edward Makumbi, Arthur Kwizera, Pauline Byakika, Willford Kirungi, Henry Kyobe Bosa, Vincent A Ssembatya, Henry Mwebesa, Diana Atwine, Jane Ruth Aceng, Yonas Tegegn Woldermariam, all members of the Uganda SARS-COV-2 Scientific Advisory Committee

## Abstract

Evidence that face masks provide effective protection against respiratory infections in the community is scarce. However, face masks are widely used by health workers as part of droplet precautions when caring for patients with respiratory infections. It would therefore be reasonable to suggest that consistent widespread use of face masks in the community could prevent further spread of the Severe Acute Respiratory Syndrome-Coronavirus 2 (SARS-CoV-2). In this study we examine public face mask wearing in Uganda where a proportion wears masks to protect against acquiring, and the other to prevent from transmitting SARS-CoV-2. The objective of this study was to determine what percentage of the population would have to wear face masks to reduce susceptibility to and infectivity of SARS-COV-2 in Uganda, keeping the basic reproduction number below unity and/or flattening the curve. We used an SEIAQRD model for the analysis. Results show that implementation of facemasks has a relatively large impact on the size of the coronavirus epidemic in Uganda. We find that the critical mask adherence is 5 per 100 when 80% wear face masks. A cost-effective analysis shows that utilizing funds to provide 1 public mask to the population has a per capita compounded cost of USD 1.34. If provision of face masks is done simultaneously with supportive care, the per capita compounded cost is USD 1.965, while for the case of only treatment and no provision of face masks costs each Ugandan USD 4.0579. We conclude that since it is hard to achieve a 100% adherence to face masks, government might consider provision of face masks in conjunction with provision of care.

## Background

A pneumonia of unknown cause detected in Wuhan, China was first reported to the World Health Organization Country Office in China on 31 December 2019 (WHO). The WHO temporarily termed the new virus 2019 novel coronavirus (2019-nCoV) on 12 January 2020. On 30 January 2020, WHO declared the outbreak a Public Health Emergency of International Concern. The WHO officially named this infectious disease coronavirus disease 2019 (SARS-COV-2) on 12 February 2020. Later, the International Committee on Taxonomy of Viruses (ICTV) officially designated the virus as severe acute respiratory syndrome-coronavirus 2 (SARS-CoV-2) based on phylogeny, taxonomy and established practice. As of June, 06 2020, 6,612,301 confirmed cases had been reported with 391,161 deaths across 213 countries and territories (WHO, Rolling updates on coronavirus disease (SARS-COV-2)).

SARS-CoV-2 is a new disease and the world is still learning about how it spreads. In general, respiratory virus infection can occur through contact (direct or indirect), droplet spray in short range transmission, or aerosol in long-range transmission (airborne transmission). With no supply of antivirals and vaccines (WHO, Coronavirus disease (SARS-COV-2) pandemic), countries and individuals are looking at other ways to reduce the spread of pandemic SARS-CoV-2, particularly options that are cost effective and relatively easy to implement. One of the preventions and controls for further spread of SARS-CoV-2 is the use of facemask, within the context of other public health interventions such as regular handwashing/sanitization, and social distancing (Chu et. al., 2020; Crowling et. al., 2020; Barasheed et. al., 2020; Lau et. al., 2020; Suess et. al., 2020; Ngonghala et. al., 2020). More recent studies have since added additional evidence of the enhanced protective value of masks (Wang et. al., 2020), and that their use would serve as an adjunctive preventive method regarding the SARS-COV-2 outbreak (Liang, Mingming et al., 2020). Masks were previously not generally recommended for the public because they can be contaminated by other people’s coughs and sneezes, or when putting them on or removing them (Kwon 2017). They may also offer a false sense of security, although they are beneficial as covers of the mouths of people already infected (WHO, Advice on the use of masks in the context of SARS-COV-2). The WHO currently recommends medical-grade mask for people over 60 years when they are out and cannot socially distance, while all others should wear a three-layer fabric mask (WHO - Coronavirus disease, 2020: Advice for the public: Myth busters). People without symptoms transmit the coronavirus without knowing they are infected (Byambasuren et. al., 2020; Wei et. al., 2020; Cheng et. al., 2020). As in prior respiratory infections (Sim, Moey, and Tan, 2014), absence of any documented community cases in Uganda appears to have led some people to perceive themselves to not be susceptible and the absence of any SARS-COV-2 related death may be perceived as a less severe infection. Recent research has shown that people are willing to wear facemasks to protect themselves against infection and further transmission for those already infected (Chu et. al., 2020). There is therefore need to quantify the effect and cost effectiveness of using facemasks in reducing the spread of SARS-CoV-2.

Uganda has adopted the use of face masks among the key interventions for prevention of the spread of SARS-COV-2, and the Government has supported expansion of local manufacturing of masks with a plan to distribute at least one free mask to all Ugandans aged 6 years and older. However, there were mixed reactions from the public with concerns about the mandatory use as well as cost of the masks. Adhering to public mask use might feel as a forfeiture of one’s freedom due to the discomfort (Scottie Andrew, CNN), while others view it as a sign of fear, weakness or vulnerability. On the other hand, initial public health recommendations did not encourage use of masks in public except for the infected individuals (WHO, 2020). These mixed messages and uncertainty of the effectiveness of masks in public, may affect compliance with use of masks. Masks are not physically comfortable (Jefferson et. al., 2020; Matusiak et. al, 2020), and this may be enough to steer some people away from them. The aim of this study is to therefore determine the efficacy and cost-effectiveness of facemasks in reducing susceptibility to and infectivity of SARS-CoV-2. We also seek to determine what percentage of the population would have to wear this gear in order to reduce the number of coronavirus infections within the community.

## Methodology

To estimate/forecast the population level impact and cost-effectiveness of public face masks, we design an *SEIAQRD* mathematical model for a population where a proportion of the general population wears face masks for primary protection against acquiring SARS-CoV-2, while the other wears masks to prevent against transmitting disease. As in other infectious diseases, we model SARS-CoV-2 using a conceptual model indicating how individuals move from the Susceptible state (at risk state) *S*, to the pre-symptomatic (those who have been exposed *E*), the Infectious (those who display symptoms after the incubation period *I*), the Asymptomatic (infected but not displaying symptoms *A)*, the institutional Quarantined *Q*(traced contacts of the confirmed positives), the recovered (*R*) and the Dead (*D*). The *SEIAQRD* is parameterized using SARS-CoV-2 data from Uganda to estimate the parameters for its transmission. We assumed 30 initial exposures where 20 are from boarder points and 10 health workers. The transmission rate is obtained by considering the probability of infection multiplied by the estimated number of contacts. To move from one state to another, we estimate the probability of transition using the duration an individual spends in a particular compartment until they are either recovered or dead.

The model attempts to determine, if facemasks are effective in reducing susceptibility to and infectivity of SARS-CoV-2, the percentage of the population that would have to wear this gear to reduce the number of coronavirus infections within the community. The analysis and figures are generated using WHO guidelines of 3.7% exposure, 10% development of symptoms, and the 80:8:8:3:1 ratio of asymptomaticity, mild, moderate, severe and critical. Initial population used is 100,000 and the number of contacts for each confirmed individual is 20. It is assumed that quarantining of contacts ends after 14 days, while isolation or viral clearance takes an average of 21 days for the asymptomatic, mild and moderate, 28 days for severe, and 35 for critical. We explored the effect of adherence to use of masks on viral reproduction, and the effect of early detection to community spread. The model was rerun with varying detection rates, when facemask adherence in both cases is 60%.

We also hypothesize that masks can reduce exposure and perform a cost-utility analysis to examine whether they are cost effective as compared to quarantining and/or treatment of symptoms. Two scenarios were considered: first, assuming that the entire population received and wore masks, we determine how much it would cost the country in comparison to the cases averted. Secondly, how many cases would be averted if the funds were invested in other mitigation measures, or in treatment? We explore this using an initial number of 30 exposed individuals in the community, 20 at boarder points and 10 health workers.

A simple model with the transit states and relative costs for each transition was used. We compare the costs for one mask to the unit costs for detection, contact tracing, quarantine, treatment including beds and ventilators. We also compare the cases averted for each intervention, and the quality adjusted life years. The stages of an infected individual are quantified using a transit state model and the unit cost for masking and treatment are compared. The cost-effectiveness analysis is quantified using the current costs by the Ministry of Health for quarantining of contacts and treatment of the symptomatic SARS-COV-2 infected individuals.

## Results

Figure 1 shows that in the case when 80% of the population do not wear masks and only 20% wear masks, one infected individual can lead to two new infections. A slight reduction in the replication number is observed, but it does not go below unity and the virus is contained. When the proportion that wears masks is increased to 80% there is a tremendous reduction in the new infections resulting from one symptomatic individual in the community (Figure 2). In Figure 2, we observe that even when modeled with a low mask coverage, the implementation of facemasks still has a relatively large impact on the size of the SARS-CoV-2 epidemic. However, the greatest reduction is observed when a high percentage wears facemask. In this case, the replication number is reduced to below unity and the virus is suppressed.

**Figure 1:**
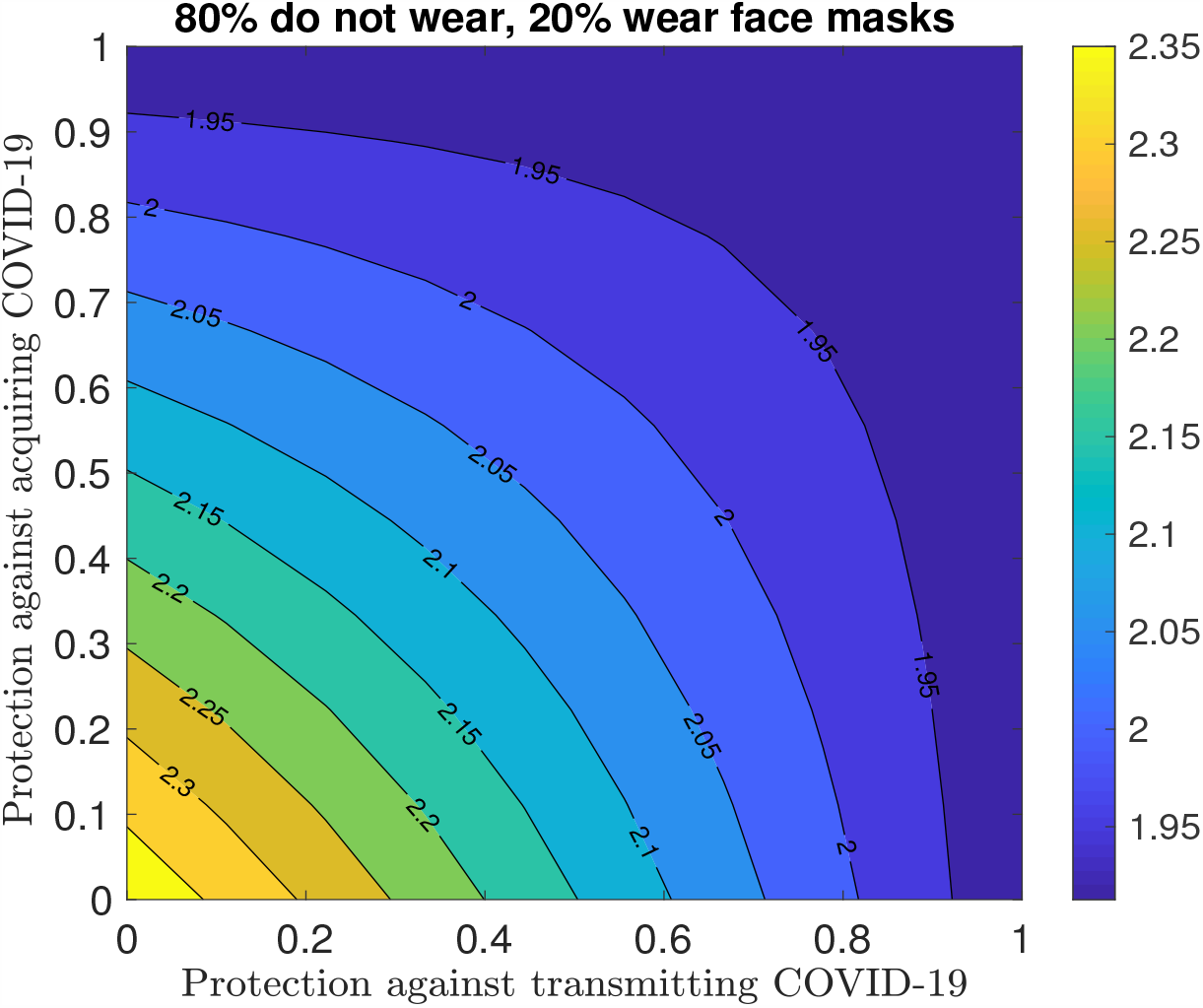
80% of the population do not wear face masks while 20% do.

**Figure 2:**
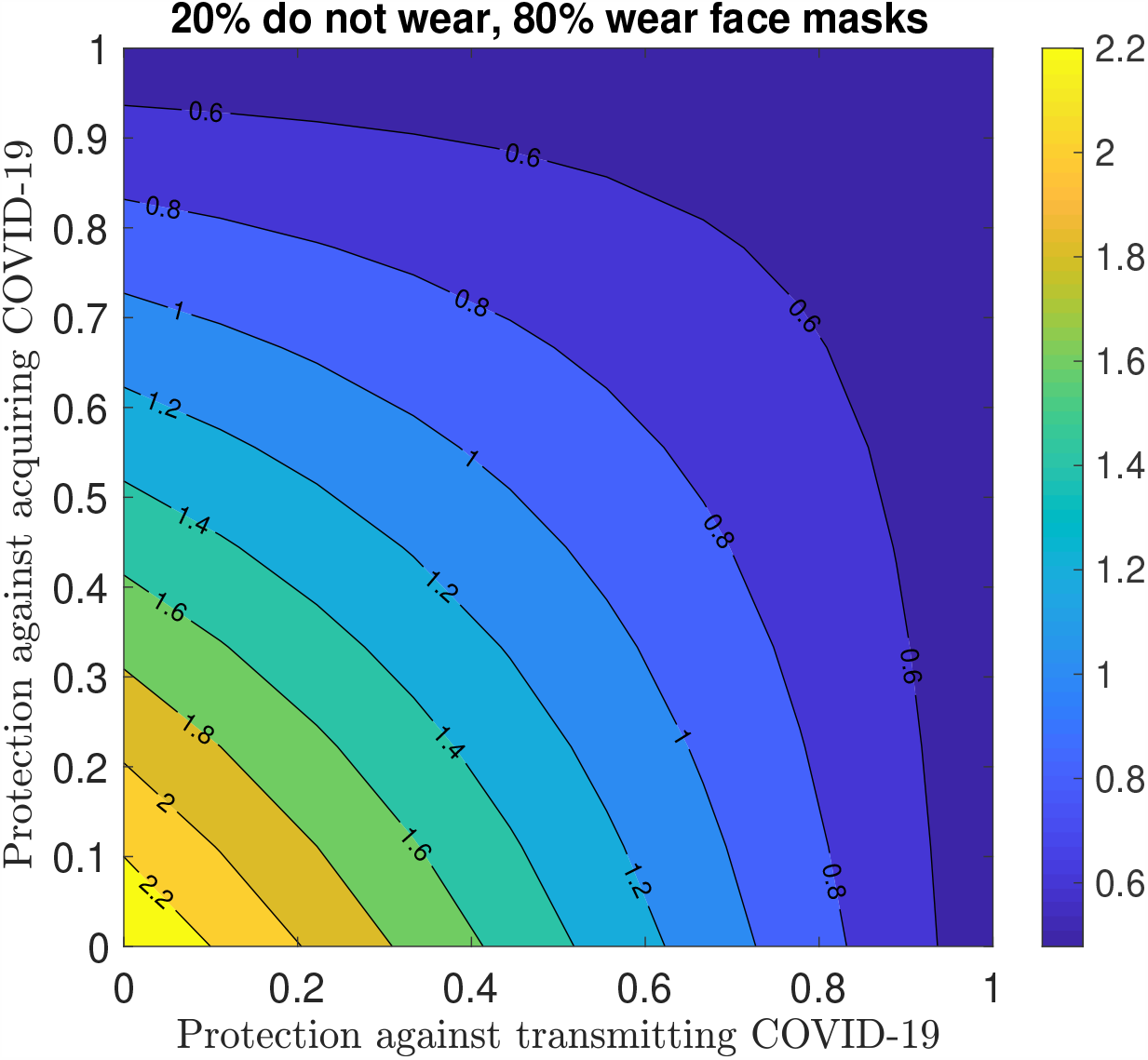
20% of the population do not wear face masks while 80% do.

Next we determine the effect of facemasks to community exposure when they are worn to protect the users. When wearing facemasks primarily to protect against exposure of the users to SARS-CoV-2, there is a high number of exposed individuals in the community. However, when facemask coverage is increased to 90-100%, there is a 34.2% reduction in exposure within 35 days (Figure 3). As seen in Figure 3, the limiting value is attained faster and this might lead to possible strong repercussions in the short run, because of limited health system capacity. When masks are worn to control the source and protect against transmitting the virus, the same reduction is observed with only 0-30% wearing the face masks (Figure 4).

**Figure 3:**
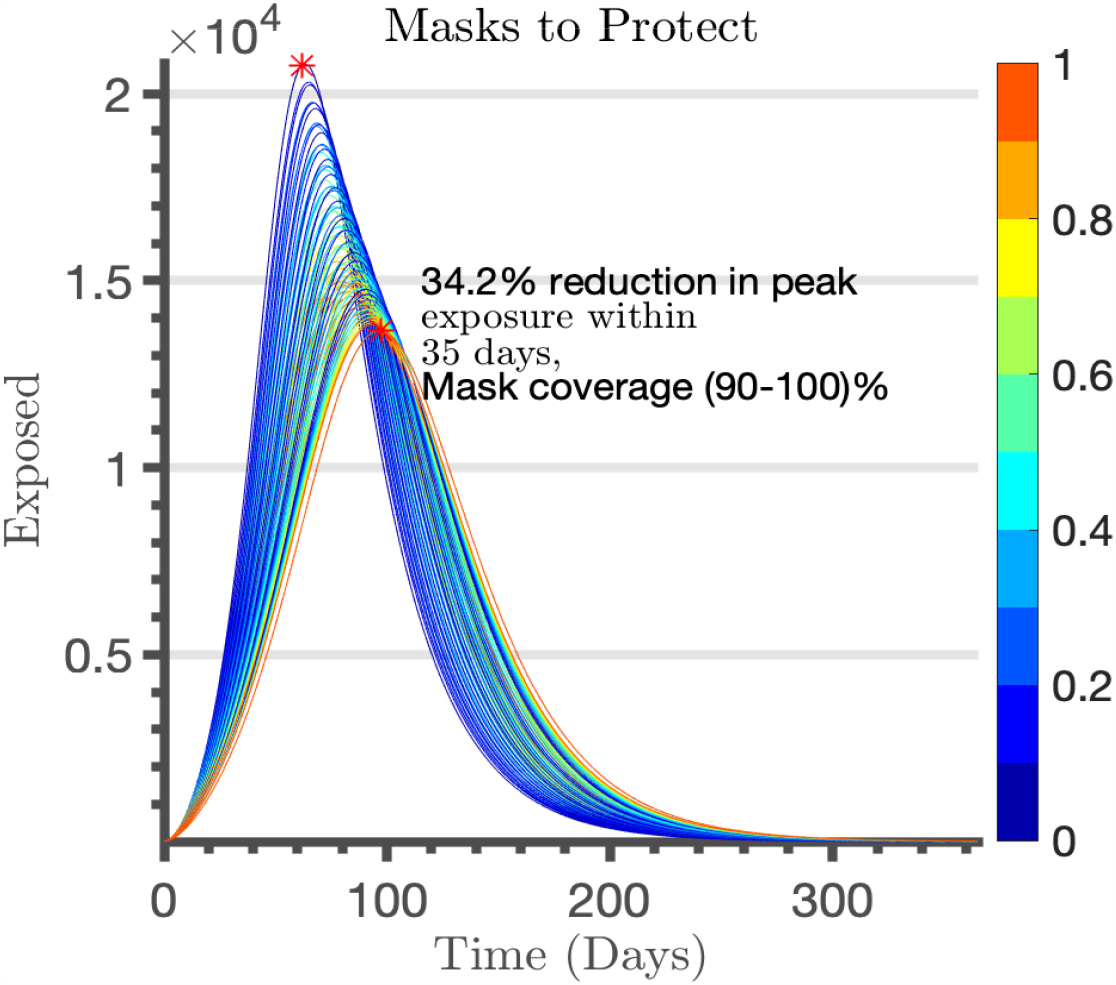
Wearing masks to reduce susceptibility.

**Figure 4:**
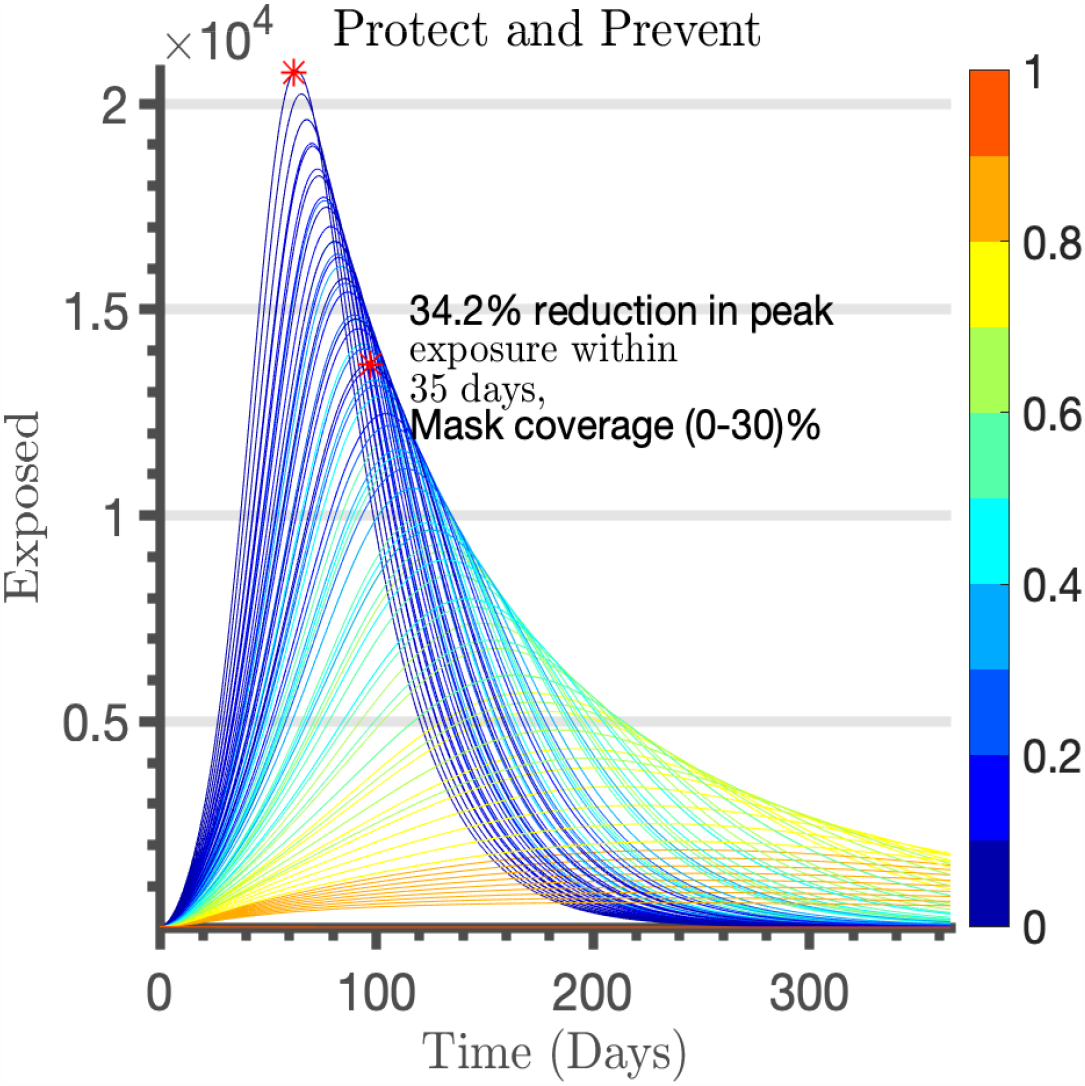
Wearing masks for source control, protection against transmitting the virus.

In this case, face masking is effective in lowering the replication number to below unity, thereby delaying the spread of the virus in the short run. When face masks are worn to protect against acquiring the virus, the replication number reduces but not below 1 and the curve is not flattened (Figure 5). In case of face masking for source control and prevention of further spread, the limiting value takes longer to be achieved but with higher reduction in exposure.

**Figure 5:**
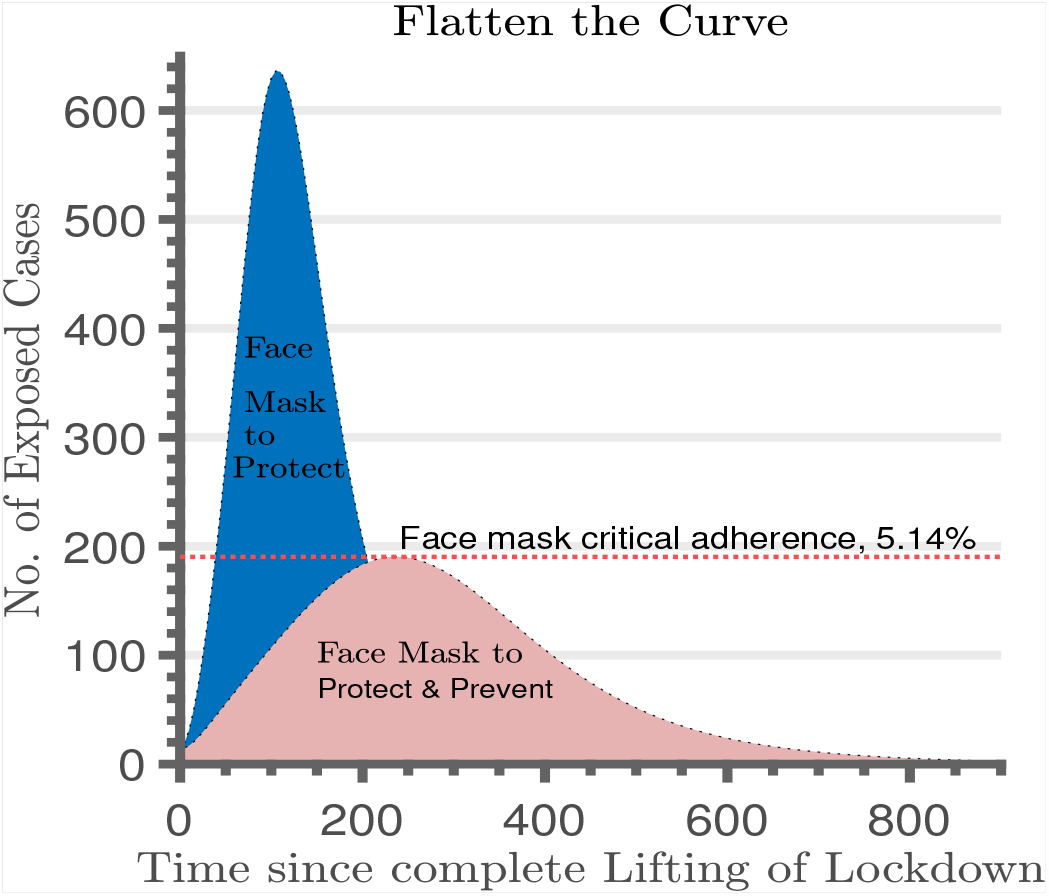
Reduction in exposure cases with public face masking.

We further observe a reduced number of cumulative deaths (42.28%) in the case of more face mask coverage (Figure 6), and extra time to expand the healthcare system capacity to provide care for those who eventually contract the virus.

**Figure 6.**
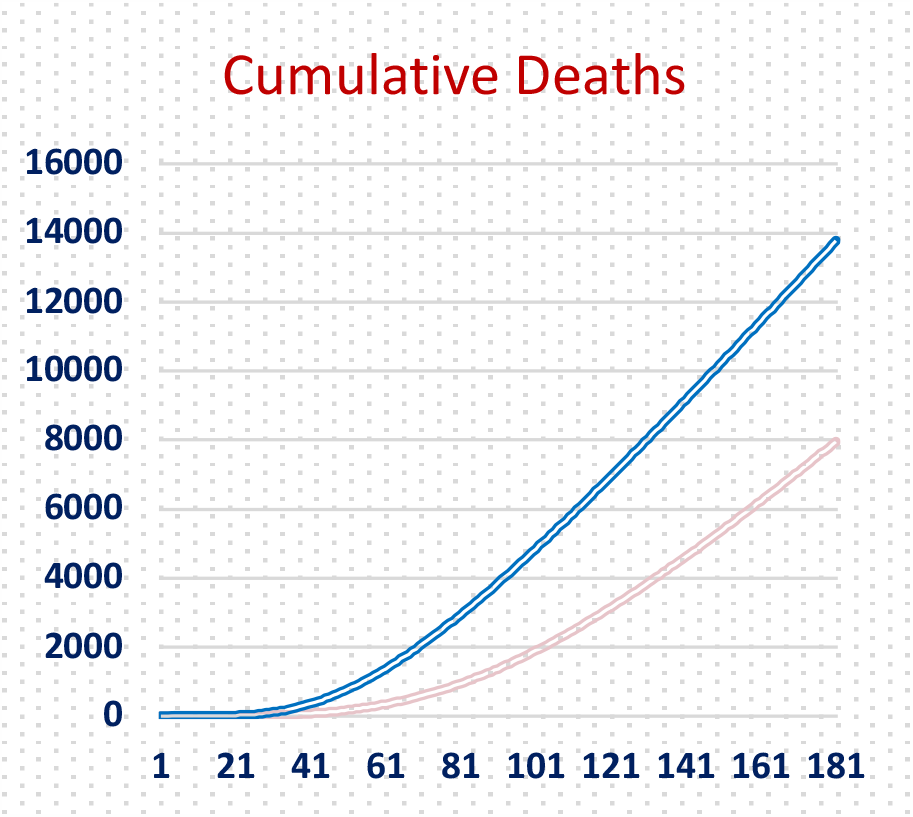
Reduction in death due to face masking.

For example, when face mask coverage is between 80-100%, there is a 91% reduction in SARS-CoV-2 exposure and the curve is flattened after 220 days. In both scenarios, compliance and adherence must be ensured. We explored the effect of adherence to use of masks on viral reproduction. We find that adherence to public facemasks as a mitigation measure reduces the replacement number to below unity, thereby controlling further spread of SARS-CoV-2 (Figure 7). We observe that when individuals adhere to face masking, it provides a faster reduction in replacement number.

**Figure 7:**
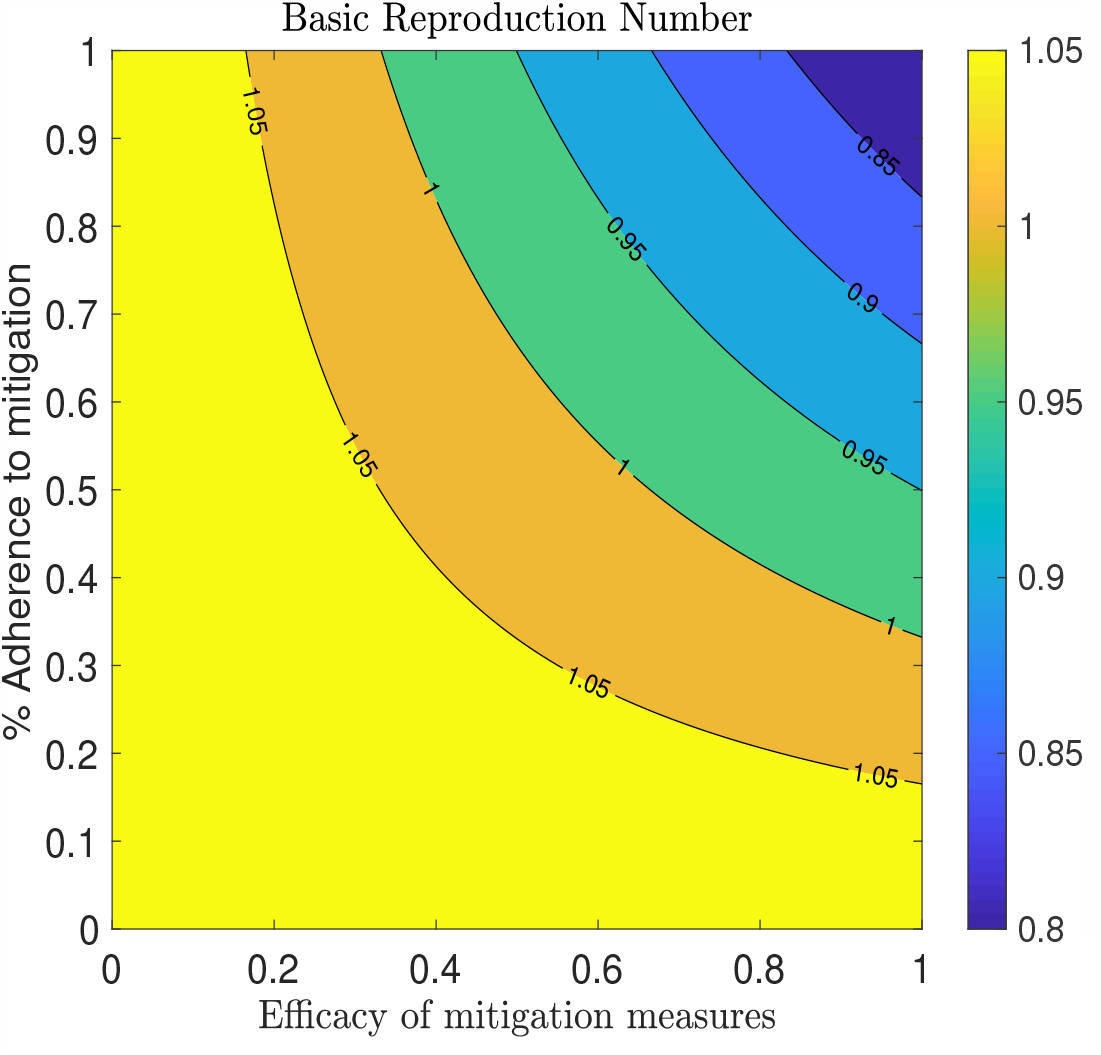
Efficacy of and effect of adherence to facemasks to disease reproduction.

We also point out that the mask used should comply with the WHO guidelines of medical-grade masks (WHO, Information on SARS-COV-2 and NCDs) or a three-layer fabric mask. With low efficiency of masks, SARS-CoV-2 will still spread within the community.

With effective public face masking, we observed a reduction in exposure. Next, we sought to determine, how detection rates affect community exposure as an additional control strategy. Currently in Uganda, once detected, an individual is quarantined until they are cured. How would the detection rate affect the number of exposed or asymptomatic individuals in the community amidst face masking?

Figures 8 and 9 show that early detection is paramount to retrieve exposed or symptomatic individuals quickly from the community to minimize further spread. As in prior cases, faster dynamics are observed when face masks are worn to reduce susceptibility, but higher reductions are obtained with high detection rates.

**Figure 8:**
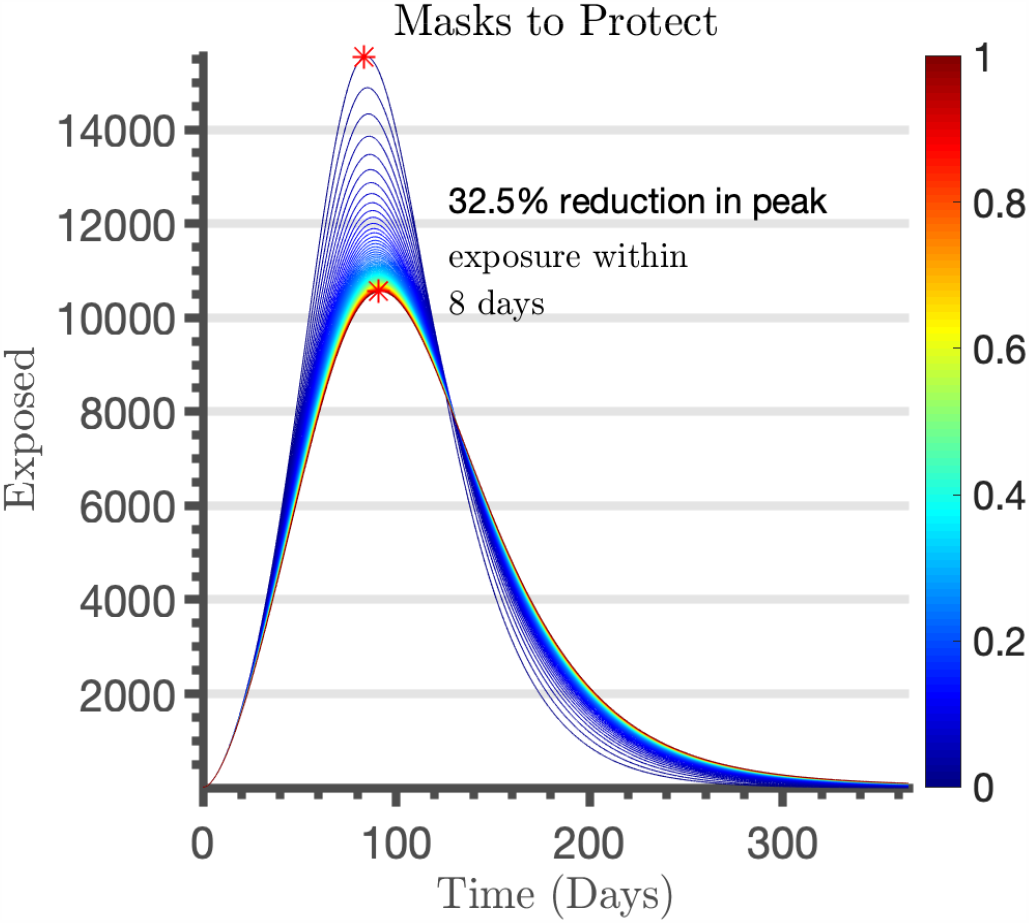
Wearing facemasks to reduce susceptibility – Effect of early detection.

**Figure 9:**
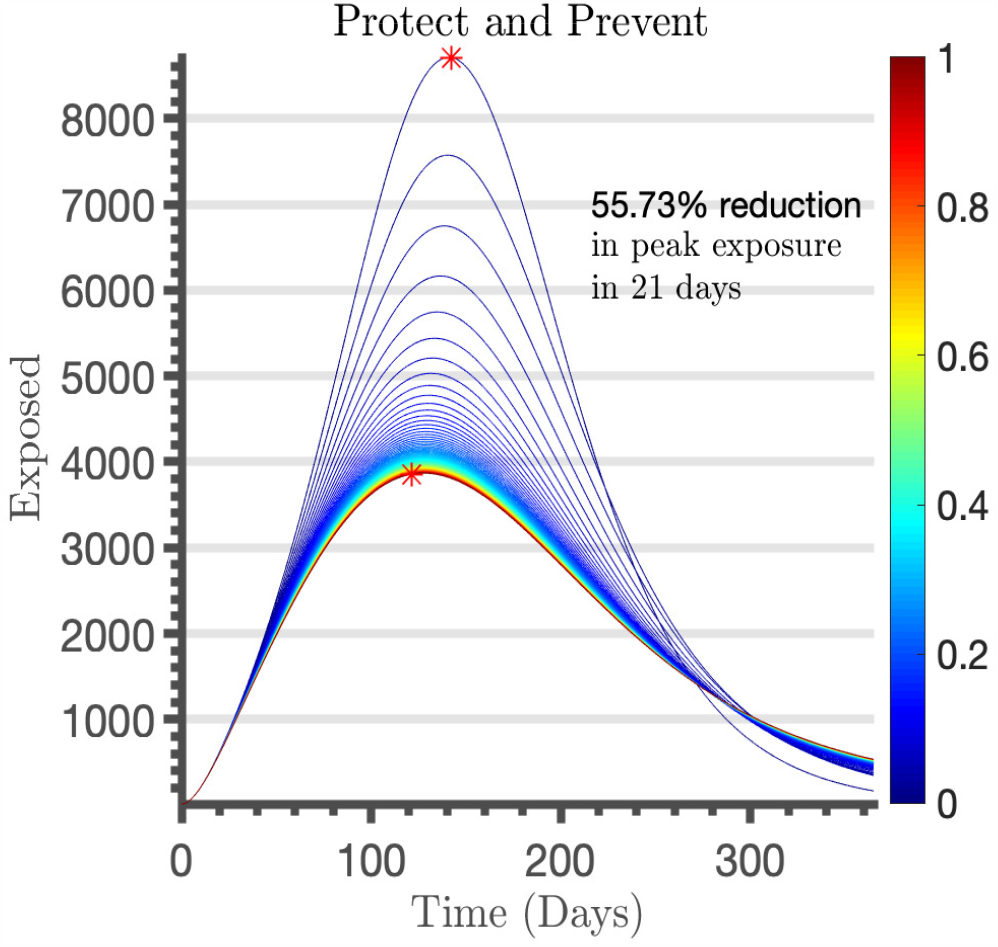
Wearing facemasks for source control, protection against transmission – Effect of early detection.

**Figure 10:**
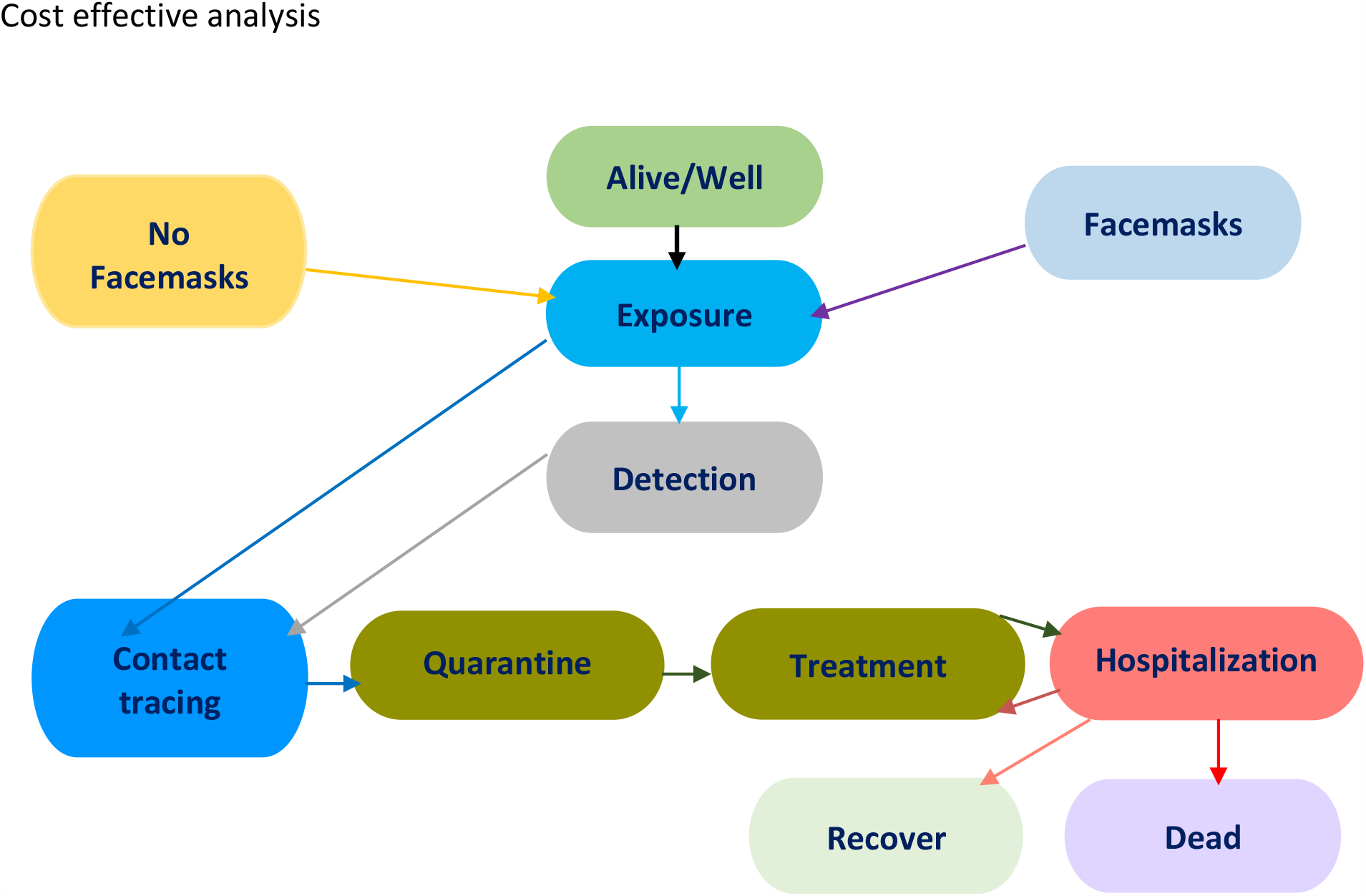
A state diagram of the model. All individuals start the model in the Alive/Well state and transition to one of face masking or not. With the exception of the dead state, suspected/confirmed SARS-COV-2 cases are transitioned between the different states

Although public face masking has been shown to be effective in controlling further spread of SARS-CoV-2, the cost-effectiveness of this strategy in Uganda is unknown. Next we determine the cost-effectiveness of facemasks in Uganda using the transit states model in Figure 9, with inputs from the Ministry of Health.

### Cost inputs

**TABLE 1:** Mitigation measures and-related costs and resources for prevention and control of SARS-COV-2 in Uganda using facemasks or treatment in a public healthcare.

| Intervention I: 100% provision of Masks of 40,000,000 |  |  |  |  |  |  |
| --- | --- | --- | --- | --- | --- | --- |
| Facemasks (40,000,000) | Cost USD/ Mask | #of Masks provided to each person (Reusable) | Total #of Masks required (Reusable) | Total Costs (USD) |  |  |
| Masks | 1.34 | 1 | 40,000,000 | 40,000,000*1*1.34 =53,600,000 |  |  |
| 1 Re-usable mask provided |  |  |  |  |  |  |
| Total Costs for providing 1 mask to 40,000,000 people |  |  |  | 53,600,000 |  |  |
| Intervention II: No masks, treatment of the infected (10% of 3.7% of 40,000,000) |  |  |  |  |  |  |
| Treatment for 3.7% Exposure, 10% successful Development of virus (163,830) | Detection/ Contact tracing/ Quarantine/Treatment |  | Cost USD/ Resource/Day | Max. Units (Days) | Total Costs | #of Individuals needing care |
|  | Asymptomatic |  | 26.89 | 21 | 564.69 | 131,064 |
|  | Mild |  | 40.23 | 21 | 844.83 | 13,106 |
|  | Moderate |  | 67.04 | 21 | 1,407.80 | 13,106 |
|  | Severe |  | 67.04 | 28 | 1,877.10 | 4,915 |
|  | Critical |  | 201.13 | 35 | 7,039.60 | 1,638 |
| Total cost for isolation facility/Hospital care (USD) |  |  |  |  |  |  |
|  | Asymptomatic |  | 74,011,000 |  |  |  |
|  | Mild |  | 11,072,000 |  |  |  |
|  | Moderate |  | 18,451,000 |  |  |  |
|  | Severe |  | 9,226,000 |  |  |  |
|  | Critical |  | 49,556,000 |  |  |  |
| Total cost For isolation facility/ Hospital care (USD) |  | 162,316,000 |  |  |  |  |

In this case we assume 30% adherence to facemasks. The total population used is 40 million, and of this, only 12,000,000 wear masks. Thus, 28,000,000 are at risk of exposure. Therefore, the number of individuals who are at risk of exposure is 3.7% of 28 million which is 1,036,000. As per WHO, only of 10% of exposures result in successful infection. Therefore, The total number of individuals who eventually get SARS-COV-2 infection is 103,600. The third intervention is providing masks for 12,000,000 and treating 103,600.

**TABLE 2:** Mitigation measures and-related costs and resources for prevention and control of SARS-COV-2 in Uganda using facemasks and treatment in a public healthcare.

| Intervention III: Masks & Treatment of the infected |  |  |  |  |  |  |
| --- | --- | --- | --- | --- | --- | --- |
| Facemasks (40,000,000) | Cost USD/ Mask | #of Masks (Reusable) | #of Masks (Reusable) | Total Costs (USD) |  |  |
| Masks | 1.34 | 1 | 12,000,000 | 12,000,000*1*1.34 =16,080,000 |  |  |
| Masks | 1.34 | 5 | 12,000,000 | 12,000,000*5*1.34 =80,400,000 |  |  |
| 1 Re-usable mask provided |  |  |  |  |  |  |
| Total Cost (USD) for providing 1 mask to 12,000,000 people |  |  |  | 16,080,000 |  |  |
| Intervention II: No masks, treatment of the infected (10% of 3.7% of 28,000,000) |  |  |  |  |  |  |
| Treatment for 3.7% Exposure, 10% successful Development of virus (103,600) | Detection/ Contact tracing/ Quarantine/Treatment |  | Cost USD/ Resource/Day | Maximum Units (Days) | Total Costs | #of Individuals needing care |
|  | Asymptomatic |  | 26.89 | 21 | 564.69 | 82,880 |
|  | Mild |  | 40.23 | 21 | 844.83 | 8,288 |
|  | Moderate |  | 67.04 | 21 | 1,407.80 | 8,288 |
|  | Severe |  | 67.04 | 28 | 1,877.10 | 3,108 |
|  | Critical |  | 201.13 | 35 | 7,039.60 | 1,036 |
| Total cost for isolation facility/Hospital care (USD) |  |  |  |  |  |  |
|  | Asymptomatic |  | 46,802,000 |  |  |  |
|  | Mild |  | 7,002,000 |  |  |  |
|  | Moderate |  | 11,668,000 |  |  |  |
|  | Severe |  | 5,834,100 |  |  |  |
|  | Critical |  | 7,293,000 |  |  |  |
| Total cost For isolation facility/ Hospital care (USD) |  | 78,599,100 |  |  |  |  |

### Summary

**TABLE 3:** Summary of costs.

| Intervention | Masks | Treatment | Masks & Treatment |
| --- | --- | --- | --- |
| Total costs (USD) | 53,600,000 | 162,316,000 | 94,679,200 |

## Discussion

In this study, we look at the efficacy of face masks as a mitigation measure against SARS-CoV-2. A mathematical model was used to determine the impact and cost effectiveness of face masks to reduce susceptibility or as a source control and prevention of transmission. Our results show that when a proper mask is used correctly, there is reduction in exposure, and a delay in the spread of the virus within the community. It also flattens the curve and provides time to expand the healthcare capacity. We caution here that these results are achievable when a proper mask is used efficiently. Any mask is only as efficient as the way it is used, fitted, worn and discarded. The reason it can cause more spreading of the virus is because people don’t fit their masks properly and continue touching the outer contaminated surface to adjust it, without hand washing or sanitizing thereafter. Therefore, our results are obtainable with continued public adherence to correct use of proper face masks. We also note that other primary interventions, such as handwashing, sanitizing and social distance “combination intervention”, are essential as public health approaches. Model simulations, using data relevant to SARS-CoV-2 dynamics in Uganda suggest that broad adoption of face masks may meaningfully reduce community transmission of SARS-CoV-2 and decrease peak hospitalizations and deaths. With these results, we caution that masks are efficient when worn to protect against catching the virus, and for source control/protection against further transmission.

Comparing the implementation costs of face masking, we find that simultaneous provision of face masks and treatment is a more efficient and perhaps sustainable approach than provision of face masks only, or provision of only treatment.

These results reinforce the current Uganda policy which integrates masks to suppress and contain SARS-CoV-2. The scenario with dual purpose to protect and prevent further transmission is more efficient and people should be made aware of this and encouraged to wear masks.

### Recommendations

This report provides an insight into the potential community-wide impact of widespread face mask use by the general population. The mathematical model parameterized using Ugandan data relevant to SARS-CoV-2 transmission suggests strong benefits of the general use of face masks in public. From the study we recommend that facemasks

i. should be nearly universal if supplies permit to ensure nation-wide compliance
ii. should be adopted as early as possible for the best population-level benefit
iii. be used concurrently with other interventions and viewed as a complement to other public health non-pharmaceutical interventions and not as an alternative use even when SARS-CoV-2 burden is low will pay dividends
iv. are most efficient for source control, but valuable as both source control and primary prevention

These are theoretical results and must be interpreted with caution, owing to a combination of potentially high rates of noncompliance with mask use in the community, uncertainty with respect to the intrinsic effectiveness of (especially homemade) masks at blocking respiratory droplets and/or aerosols, and uncertainty regarding the basic mechanisms for respiratory infection transmission. However, these results suggest a significant value even to low quality masks when used widely in the community. Despite the uncertainty, the potential for benefit, the lack of obvious harm, and the precautionary principle, we strongly recommend facemask use nationwide, especially if there is no diversion for healthcare supply.

## Data Availability

Data available upon request

## Acknowledgements

The authors acknowledge all members of the Uganda SARS-COV-2 Scientific Advisory Committee. We further acknowledge the Centers for Disease Control and Prevention Uganda partnership and contribution.

